# Higher Body Mass Index is an Important Risk factor in COVID-19 patients: A Systematic Review

**DOI:** 10.1101/2020.05.11.20098806

**Authors:** Vivek Singh Malik, Khaiwal Ravindra, Savita Verma Attri, Sanjay Kumar Bhadada, Meenu Singh

## Abstract

**Background:** Globally, both obesity and underweight are severe health risks for various diseases. The current study systematically examines the emerging evidence to identify an association between Body Mass Index (BMI) and COVID-19 disease outcome.

**Methods:** Online literature databases (e.g., Google Scholar, PubMed, MEDLINE, EMBASE, Scopus, Medrixv and BioRixv) were screened following standard search strategy having the appropriate keyword such as “Obesity”, “Underweight”, “BMI”, “Body Mass Index”, “2019-nCov”, “COVID-19, “novel coronavirus”, “coronavirus disease”. Studies published till 20^th^ April 2020 were included without language restriction. These studies include case reports, case series, cohort, and any other which reported BMI, overweight/obesity or underweight, and its complication with COVID-19 disease.

**Findings:** Obesity plays a significant part in the pathogenesis of COVID-19 patients, though the role of BMI in the COVID-19 pandemic must not be ignored.

**Interpretation:** Consequences of inflammation of adipose tissue has been reported as a leading cause of insulin resistance and hypertension due to metabolic dysfunction. The results of the current study show that BMI plays a significant role in COVID-19 severity in all ages, especially the elderly population. A panel should review COVID-19 patients with higher BMI and other co-morbidities, and they should be given increased vigilance, testing priority, and therapy. Further, the COVID-19 patients whose illness entered 7-10 days, age >50 yrs, and elevated CRP levels should have additional medical considerations.

**Recommendation:** Population and patients with high BMI have moderate to high risk of medical complications with COVID-19, and hence their health status should be monitored more frequently.

## Introduction

Being overweight, obese, and underweight may have a major risk factor for multiple disorders in the later stage of life. Earlier studies have shown a relationship between BMI and mortality among diabetic patients (1) mental behavioral, neurological (2) Parkinson’s disease (3), weight gain with pneumonia (4). Infection was reduced in weight-reduced patients (5), and thromboembolic risk in obese (6).

### Influence of infection on obesity

An U-shaped elevated infection rate was seen among overweight/obese and underweight old age persons (7), increased CVD risk with greater malignancy rates (8–10), and obesity with impaired immune responses was seen (11). overweight/obesity was reported as a risk factor for Clostridium difficile colitis, pneumonia, bacteremia infections (12,13), and increased surgical site infection (14–20). Similarly, Waist circumference was reported as a better predictor of septicemia risk than BMI (21). In a study, Adipose tissue serves as a source for human influenza A virus, Trypanosoma gondii, HIV, adenovirus Ad-36, cytomegalovirus, and mycobacterium tuberculosis (6). The elderly population is reported to be more susceptible to infections (22). Higher mortality risks of influenza was reported among the underweight and obese old population (23).

## Obesity and Coronavirus Infection

In many countries, the obese population is more vulnerable to most of the non-communicable diseases. Further, the COVID-19 pandemic has put this population at higher risk (24). The measures introduced in some countries, e.g., restriction on leaving home for several weeks even for daily-walk, will have an impact on mobility, and these infection control measures resulted in physical inactivity, and even short periods of restriction can increase the risk of metabolic disease in future. Overweight and obesity among severe COVID-19 patients were reported as an independent risk factor (25). Increased adiposity undermines the pulmonary function contributing the viral pathogenesis as a secondary cause of infection (26) and has been reported as evidence in obese among older (>60 years of age) (27). Following the 2009 influenza A virus H1N1 pandemic (28) and Adult Respiratory Distress Syndrome (ARDS) (29), obesity was reported as a risk factor. Decreased mortality due to ventilator-induced lung injury resulting in chronic pro-inflammatory status in obese patients has been reported (30). Infection of influenza A showed that overweight/obesity risk elevates with virus shedding (42% longer) duration among symptomatic patients with obesity (31). In H_1_N_1_ disease, obesity was reported as a risk factor with increased hospitalization and death rates (32). Obesity has been shown to increase vulnerability to infections, serving as a risk factor among COVID-19 patient’s mortality rate (33).

### Potential Confounders for obesity and infection risk

Smoking behavior (4,34), well-being (35), Physical activity (36), weight loss (7), Co-morbidities (37), and nutrition (38) are potential confounders reported independent of age and sex.

##### Research in context

###### Evidence before this study

We searched PubMed for and other online databses for reported overweight/obesity or BMI and COVID-19. In case of COVID-19, a toal 14 studies have been reported and included for systematic review, for meta-analysis only 10 studies have been added.

###### Added value of this study

This systematic review and meta-analysis addressing the role of higher BMI among COVID-19 patients. We systematically reviewed PubMed, EMBASE, Medrixv and BioRixv) using relevant keywords.

###### Imlications of all the avialeble evidence

Population and patient with high BMI has moderate to high risk of medical complication with COVID-19, and hence their health status should be monitored more frequently.

## Methods

### Search Strategy

PRISMA guideline was used for this meta-analysis (39). **Boolean operators:** (BMI) AND (COVID-19), (Obesity) AND (COVID-19), ((Underweight) AND (COVID-19)) OR (CASRS-Cov-2), (BMI) OR (Body Mass Index) AND (COVID-19) were used for PubMed database and for Google Scholar, MEDLINE, EMBASE, Scopus, Medrixv and BioRixv using appropriate keywords (e.g., “Obesity”, “BMI”, “Body Mass Index”, “2019-nCov”, “COVID-19, “novel coronavirus”, “coronavirus disease”). The study included published literature without language restriction until 20^th^ April 2020.

### Selection criteria (Inclusion/exclusion)

Studies with the following conditions had included for the meta-analysis. (1) Case report, case series and cohort studies design (2) BMI assessment >25 kg/m^2^ and <25 kg/m^2^ reported (3) indicating the risk ratio and/or odds ratio for the obesity risk (4) studies reporting cross-sectional were excluded (5) age and gender were not kept as a bar for inclusion (6) For meta-analysis we included BMI with reported COVID-19 infections (clinical, laboratory or both confirmed) (7) Definition of severe COVID-19 was taken (40).

### Data extraction

Details of authors, total sample size, and numbers reported for obesity and other conditions, e.g., comorbidity (obesity) and clinical condition (critical/severe) were extracted and recorded independently. Data extraction was accomplished by two reviewers (Dr. Khaiwal Ravindra & Dr. Neha Chanana). Any disagreement was resolved by joint discussion. To minimize the risk of duplication of data carefully handled. Continuous variable (BMI) was expressed as mean ± SD/median and IQR (interquartile range). The systematic include case reports, case series and observational and any other study types of study design which reported obesity and or underweight or its complication of COVID-19 infection. Studies for meta-analysis studies were pooled only if the outcome measured in the same way by all studies.

### Quality assessment

The Newcastle Ottawa scale for cohort studies was used for quality evaluation of the selected studies (41).

### Statistical analysis and data synthesis

After extracting the results of the studies, were pooled and the effects of BMI on COVID-19 patients were examined using the random effects method. For continuous outcome standard error (SE) with 95% CI was calculated. The heterogeneity (*I^2^* statistic) was assessed between studies. Higgins and colleagues suggested *I^2^* values 25% (low), 50% (moderate), and 75% (high) indicating the existence of heterogeneity (42) (43,44). Data analysis was undertaken using Microsoft Excel (45).

## Results

### Literature search

Literature searching and screening was done according to PRISMA chart, as shown in figure 1 (supplementary). Initially, 3405 published research articles were identified through a database search. After the removal of 3368 publications due to duplicates and not relevant to study criteria, only 37 research papers were taken for the full-text paper. Finally, 14 articles met the inclusion criteria were included in the quantitative synthesis of the current systematic review.

**Figure 1:**
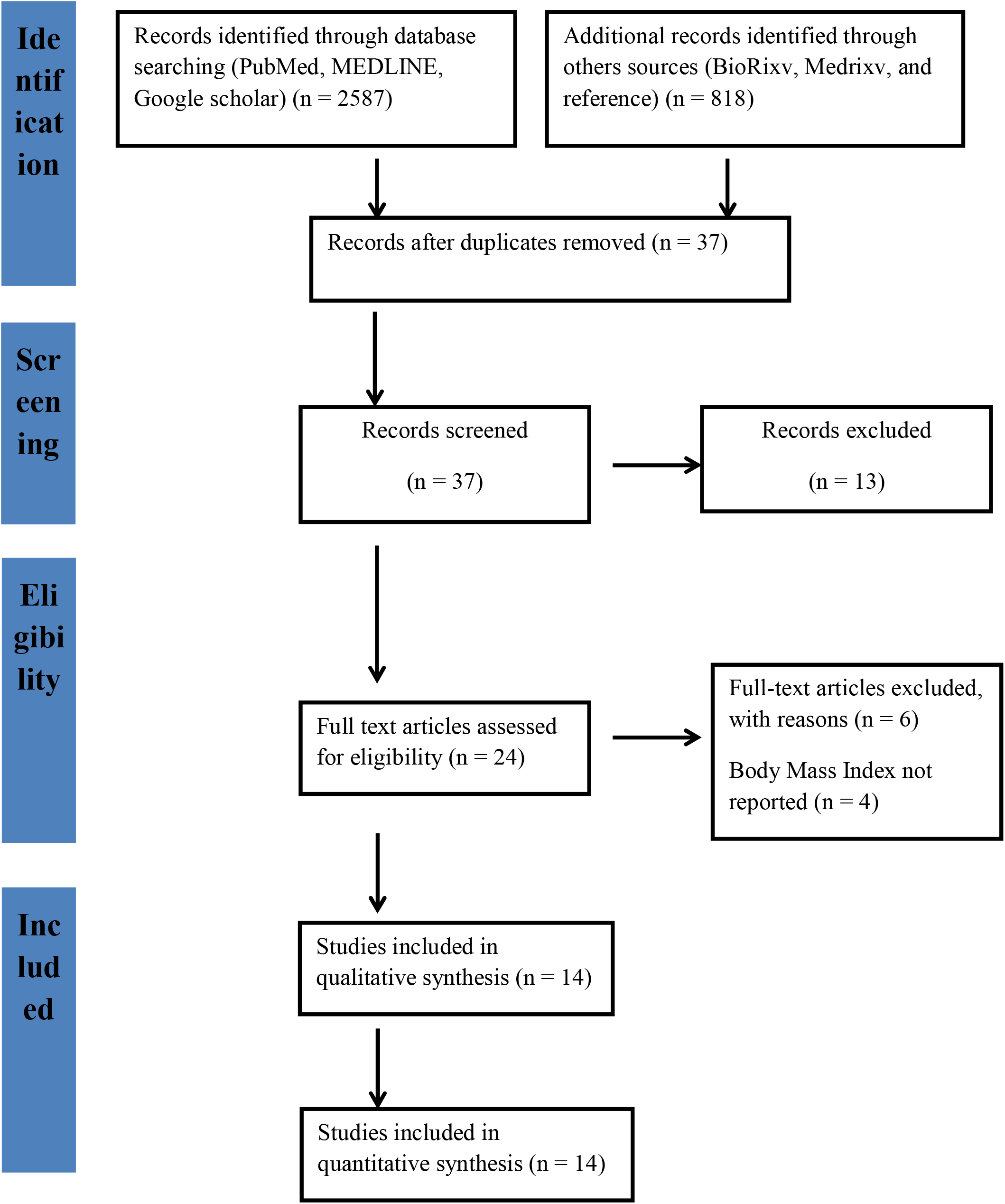
PRISMA Chart

### Study characteristics

The main characteristics were summarized in table 1(supplementary). All published research papers fall under observational (cohort) study design. Most of the studies are from China, the USA, and France. The study included articles published/available online till 20^th^ April 2020. About the obesity criteria, BMI >25 kg/m^2^ was considered as also described in selected studies.

**Table 1:**
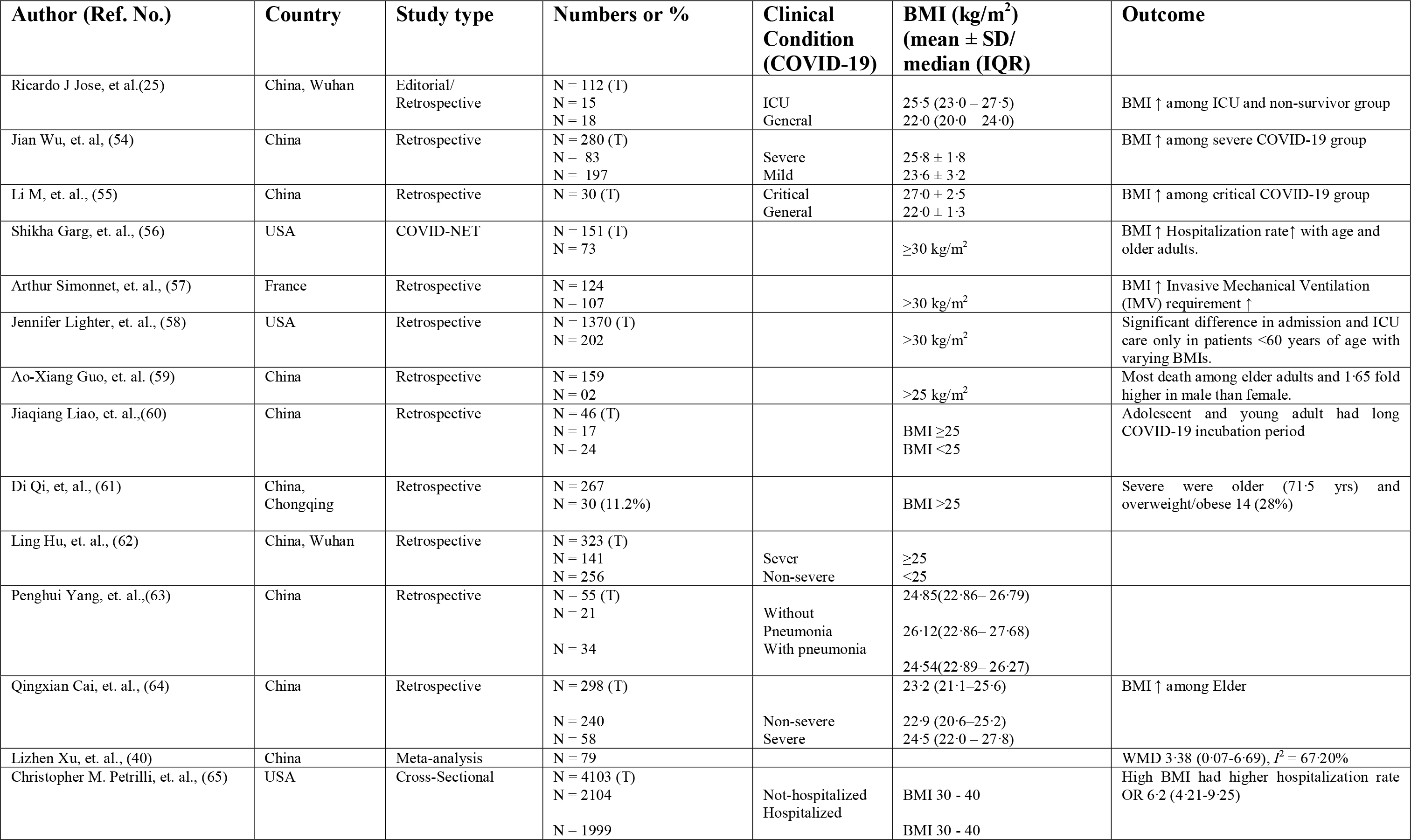
Patient/study characteristics

### Quality assessment

The Newcastle Ottawa Scale (for cohort studies) was used for the qualitative evaluation of the included studies (41). The risk of bias was assessed based on three domains, i.e., selection, comparability, and outcome, as highlighted in Table 3 (supplementary).

### Publication bias

This study includes published studies, as well as unpublished literature on MedRxiv and Bioxiv, as long as it meets study inclusion criteria. Possible publication bias was not calculated due to the limited power among studies, and the outcome was <10 for funnel plot (46).

### Meta-analysis

The outcomes of the meta-analysis (Table 2) and forest-plot (Figure 3) are shown. A random-effects model (*I*^2^ = 53·45%) was used on 9 articles for those reported ≥25BMI kg/m^2^ for COVID-19 infection. involving 689 patients to analyse the risk factors of ≥BMI kg/m^2^ [RR, 95%CI: 0·34 (0·23-0·44)] for patients with COVID-19 than those of having <25BMI kg/m^2^ (RR, 95%CI: 0·60 (0·34-0·86)] with random-effect model (*I*^2^ = 0%). BMI <25 kg/m^2^ random effect-models *I*^2^ were negative, which indicates no observed heterogeneity for this group (47), (48).

**Table 2:**
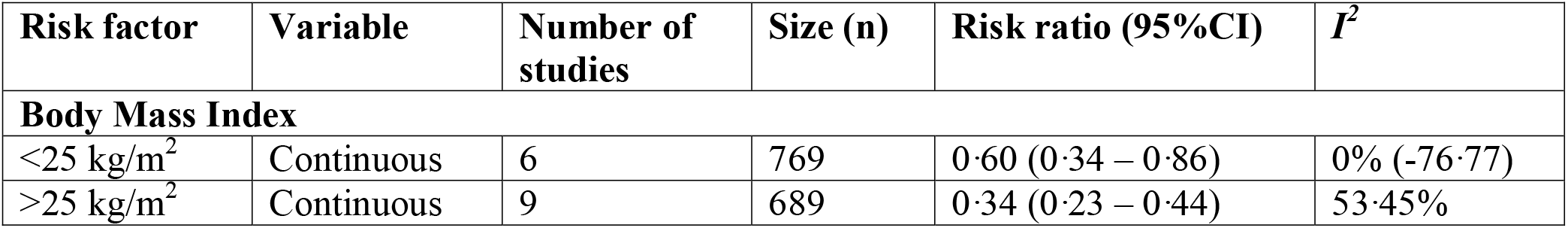
The meta-analysis of Body Mass Index for COVID-19 Patient’s

**Table 3:** Quality assessment: Cohort study Quality according to Newcastle-Ottawa Scale.

| S.No. | Selection***** |  |  |  | Comparability** | Outcome***** |  |  | Total quality score |
| --- | --- | --- | --- | --- | --- | --- | --- | --- | --- |
|  | 1 | 2 | 3 | 4 | 5 | 6 | 7 | 8 |  |
| Ricardo J Jose, et al.(25) | * | 0 | * | 0 | 0 | ** | 0 | 0 | 3 |
| Jian Wu, et. al, (54) | * | 0 | * | 0 | * | ** | 0 | 0 | 4 |
| Shikha Garg, et. al., (56) | * | 0 | * | 0 | 0 | ** | 0 | 0 | 3 |
| Arthur Simonnet, et. al., (57) | * | 0 | * | * | * | ** | 0 | 0 | 5 |
| Jennifer Lighter, et. al., (58) | * | 0 | * | * | 0 | ** | 0 | 0 | 4 |
| Jiaqiang Liao, et. al.,(60) | * | 0 | * | * | 0 | 0 | 0 | 0 | 3 |
| Di Qi, et, al., (61) | * | 0 | * | 0 | 0 | ** | * | 0 | 4 |
| Ling Hu, et. al., (62) | * | 0 | * | 0 | * | ** | 0 | 0 | 4 |
| Penghui Yang, et. al.,(63) | * | 0 | * | 0 | * | ** | * | 0 | 5 |
| Qingxian Cai, et. al., (64) | * | 0 | * | 0 | * | ** | 0 | 0 | 4 |

**Note: Selection;** 1) Representativeness of the exposed cohort, 2) Selection of the non-exposed cohort, 3) Ascertain exposure, 4) Demonstration that outcome of interest was not resented at stat of study; **Comparability;** 5) Comparability of cohorts on the basis of the design or analysis controlled for confounders; **Outcome:** 6) Assessment of outcome, 7) Was follow-up long enough for outcomes to occur, 8) Adequacy of follow-up of cohorts.

**Figure 2:**
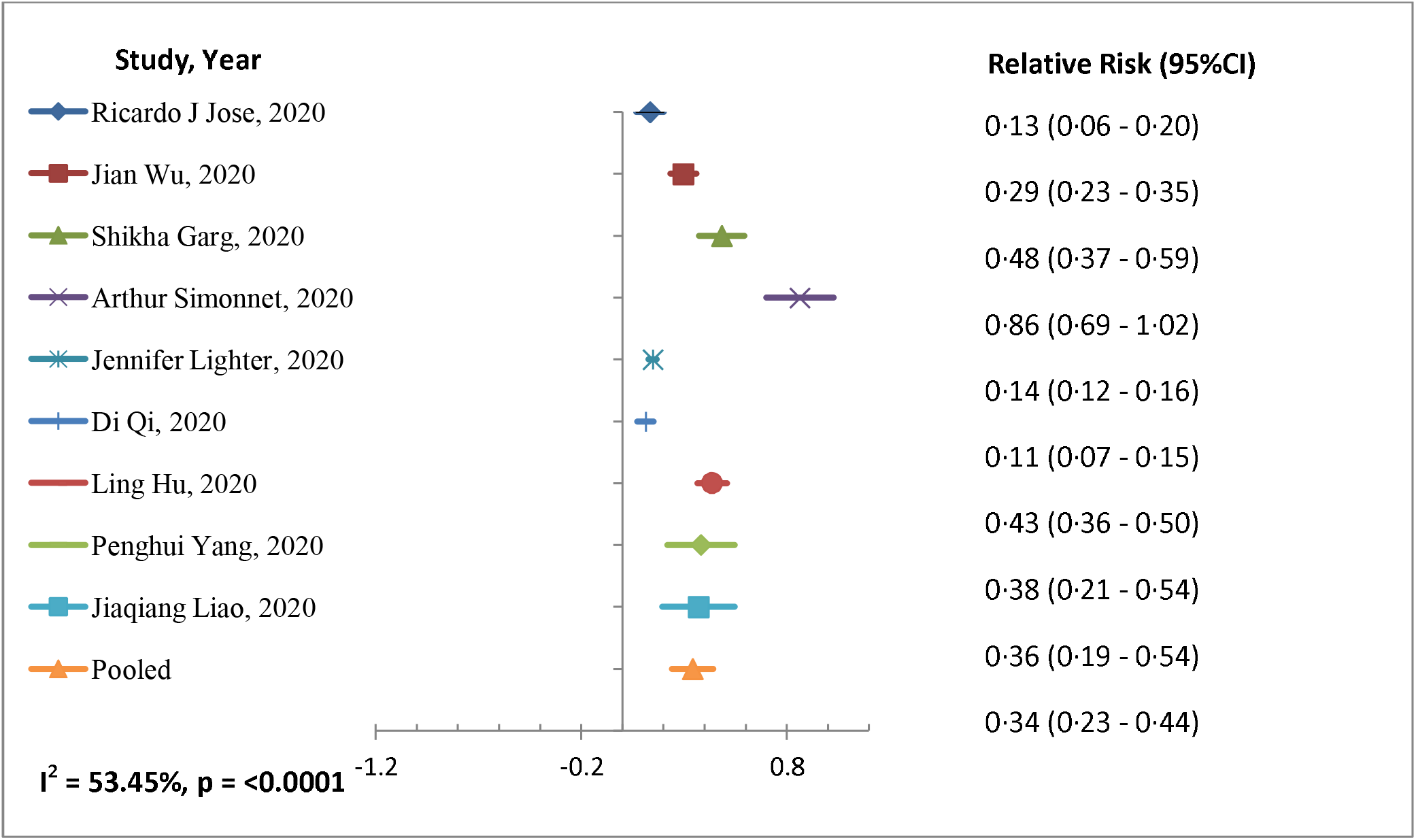
Forest plot of risk factor BMI >25 kg/m^2^ with COVID-19 patient’s

**Figure 3:**
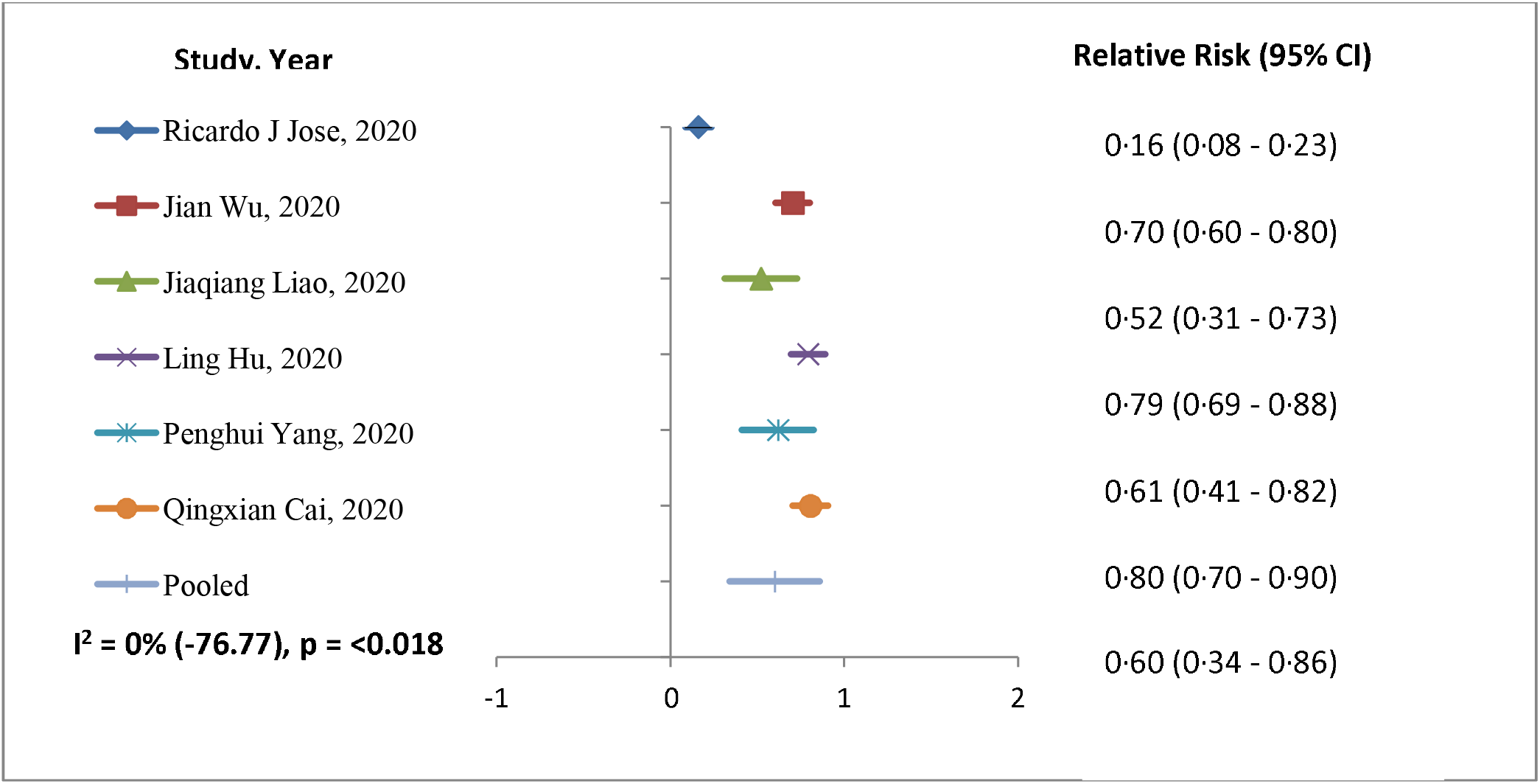
Forest plot of risk factor BMI <25 kg/m2 with COVID-19 patient’s

## Discussion

There is geographical variation in fatality case rates in South Korea (0·8), China (2·3), and Italy (7·2) has been reported (49) with risk factors of smoking, pollution, and aging. In US patients <60 years were 2 (95%CI: 1·6 – 2·6) and with BMI 30-40 (OR (95%CI)1·8: 1·2 – 2·7) more likely to be admitted to acute and critical care to individuals with a BMI <30 (50) has been reported for the first time. In a study, gender (male), age, and heart disease were the main risk factors of COVID-19 related death (51). Adolescents and young adults might play a key role in the worldwide spread of COVID-19 disease because they study overseas and frequently travel (52). Meta-analysis showed elderly male patients with high BMI have greater chances of being into critically ill patients category. (53)

### Limitations

Though there are limitation in the current systematic review and meta-analysis. Population, continuous variable, clinical condition, and statistical methods have the potency to differ and may cause heterogeneity among studies included for the meta-analysis. Further, the study reviewed only the risk of BMI (>25 kg/m^2^ and <25 kg/m^2^) of COVID-19 subjects and their severity.

### Study Importance

This is the first study with a large number of studies, linking BMI a critical risk factor for COVID-19 infection and severity. Hence, while treating, COVID-19 patients with higher BMI should be given special medical consideration as they also have other comorbidities.

### Implications of our study

This is the first meta-analysis to give an account of the higher BMI and COVID-19 infection.

### Conclusion

The BMI play aa significant role in COVID-19 infection and severity in all ages, especially elderly a population. COVID-19 patients with higher BMI should be reviewed by a panel for the risk factors. Further, there should be a procedure for increased vigilance, testing a priority, and therapy for patients with obesity and COVID-19 disease whose illness has entered 7-10 days, having age >50 yrs, and elevated CRP levels. The severity of COVID-19 has to be found a significant burden on intensive care resources in hospitals worldwide and specifically in lower-and-middle income countries due to lack of health finance and resources. Hence, patients with higher BMI with other comorbidities should be given special consideration to avoid the severity of the disease and morbidity.

## Data Availability

this is an systematic review and meta-anlysis

## Acknowledgment

We thank the Department of Community Medicine & School Public Health and Indian Council of Medical Research, New Delhi. We also thanks Dr. Neha Chanana for doing an independent review of studies included in the current systematic review and meta-analysis.

